# The Impact of Estrogen-Suppressing Contraceptives on Behavioral and Functional Difficulties in Borderline Personality Disorder

**DOI:** 10.1101/2024.09.04.24313069

**Authors:** Seyma Katrinli, Alex O. Rothbaum, Raneeka DeMoss, William C. Turner, Ben Hunter, Abigail Powers, Vasiliki Michopoulos, Alicia K. Smith

## Abstract

Borderline Personality Disorder (BPD) is characterized by rapidly shifting emotional, interpersonal, and behavioral symptoms, and is often co-morbid with mood and anxiety disorders. Females are more likely to be diagnosed with BPD than males and exhibit greater functional impairment. Hormonal fluctuations, particularly in estrogen levels, may influence the manifestation of BPD symptoms. Here we investigated the influence of estrogen-suppressing contraceptives on behavioral and functional difficulties in BPD. The analytical sample included 348 females ages 18-50 undergoing residential treatment for psychiatric disorders, with 131 having a BPD diagnosis. Patients were categorized based on their contraceptive method: 1) Estrogen-suppressing contraceptives (N=145) and 2) Naturally cycling (N=203). Interaction models tested the impact of estrogen-suppressing contraceptives on the relationship between BPD diagnosis and behavioral and functional difficulties at admission and discharge, assessed by the four Behavior and Symptom Identification Scale (BASIS-32) domains: difficulties in relationships, daily living, depression/anxiety, and impulsivity. Females with a BPD diagnosis were more likely to use estrogen-suppressing contraceptives compared to those without BPD (p=0.04). However, estrogen-suppressing contraceptive use was not associated with behavioral and functional difficulties at admission, discharge, or over time. Estrogen-suppressing contraceptives moderated the association between BPD diagnosis and difficulties in relationships (p=0.004), difficulties in daily living (p=0.01), and depression/anxiety symptoms (p=0.004). Patients with BPD expressed increased behavioral and functional difficulties at admission, discharge, and over time only if naturally cycling (p<0.003). Our findings suggest that estrogen-suppressing contraceptives may help to regulate the rapidly shifting emotional, interpersonal, and behavioral symptoms in females with BPD by stabilizing estrogen levels.

## Introduction

Borderline personality disorder (BPD) is a severe psychiatric condition that affects 6% of the adult population. The prevalence of BPD reaches up to 10% in psychiatric outpatients and 20% in psychiatric inpatients (1). Individuals with BPD suffer from rapidly shifting emotional, interpersonal, and behavioral symptoms, including unstable and chaotic interpersonal relationships, intense and highly variable emotions, impulsive behavior, aggressiveness, and chronic suicidality and self-harm behaviors (2). Due to the rapid shifts in symptoms, BPD is difficult to diagnose, and its symptoms are challenging to manage (2). Indeed, BPD is highly co-morbid with other psychiatric disorders, including major depressive disorder (MDD) and post-traumatic stress disorder (PTSD), with over 70% of individuals with BPD meeting the criteria for other psychiatric diagnoses (3).

BPD is predominantly diagnosed in females and displays distinct differences in symptoms and co-morbidities between sexes (4). Males with BPD are more likely to exhibit aggressiveness and impulsivity, often co-morbid with substance use disorders. In contrast, females tend to show emotional instability, suicidal or self-harm behaviors, and unstable relationships, along with comorbid anxiety and mood disorders (4). The biological mechanism behind these rapid shifts in symptoms may be linked to ovarian hormones, given their fluctuations across a female’s lifespan—puberty, pregnancy, menopause—and even within the menstrual cycle. Indeed, growing evidence suggests that fluctuations in ovarian hormones play a significant part in the increased prevalence of MDD, PTSD, and BPD, in females (2, 5–10). Specifically, within-person variations but not absolute levels of estrogen correlated with higher BPD symptoms (11). Moreover, individuals who exhibited higher BPD traits tended to experience increased symptoms and negative and interpersonal emotions (i.e., anxiety, rejection sensitivity, irritability, and aggression) when their estrogen levels were below average and progesterone levels were above average, a pattern typically seen during the mid-luteal and perimenstrual phases (8–10).

Given the impact of fluctuating estrogen levels on mental health symptoms, contraceptives that suppress ovulation and stabilize estrogen levels (i.e., estrogen-suppressing contraceptives) may impact the manifestation and severity of symptoms related to BPD. However, research examining whether estrogen-suppressing contraceptives improve or worsen mental health symptoms is conflicting (12–16). Rather than a straightforward association, the recent evidence suggests a complex relationship between estrogen-suppressing contraceptives and mental health outcomes depending on psychiatric history (14). Specifically, while hormonal contraceptives were associated with decreased depressive symptoms in women with a history of psychiatric disorders, in women without a history of psychiatric disorders, hormonal contraceptives increased the risk of depression (14). However, these studies focused on symptoms of depression and anxiety, which may have different underlying mechanisms than BPD (12–15). Despite multiple studies establishing the link between fluctuating ovarian hormones and BPD symptoms [11, 12, 14], only one small study to date examined the role of oral contraceptive pills, but not other hormonal contraceptives, in BPD symptoms [13]. In this study of 17 females, the use of oral contraceptive pills worsened BPD symptoms [13]. To delineate the broader impact of estrogen-suppressing contraceptives in BPD, we sought to investigate how estrogen-suppressing contraceptives influence behavioral and functional difficulties in females with and without a BPD diagnosis.

## Methods

### Participants

The participants were patients who were admitted to a non-profit residential and ambulatory psychiatric rehabilitation facility in the southeastern United States. We conducted a medical review of 413 non-menopausal and non-pregnant patients with female sex assigned at birth, ages 18 – 50, who received residential treatment for between 01/01/2019 and 02/14/2024. The average duration of treatment was 90 days. All participants have a primary diagnosis of a mood disorder, anxiety disorder, psychotic disorder, or neurodevelopmental disorder. The study was approved by the Institutional Review Board of Emory University.

### Measures

All demographic, clinical, and medical information, including psychiatric diagnoses, assessments, and contraception methods, were abstracted from medical records from the intake visit and initial history and physical. BPD diagnosis was determined using the DSM-5 criteria (1) during clinical interviews with the admitting psychiatrists in the psychiatric treatment program. Behavioral and functional difficulties were assessed at admission and discharge by the four subscales of the 32-item Behavior and Symptom Identification Scale (BASIS-32), which are difficulties in relationships, difficulties in daily living, depression and anxiety, and impulsivity (17). The BASIS-32 measures the severity and impact of various symptoms and functional problems associated with psychiatric disorders (17).

Contraception methods were abstracted from patients’ current medications in their medical records. Eligible participants were on contraception at the time of admission to the residential treatment, and they stayed on contraception during their treatment. All medication, including contraceptives, are distributed by nursing staff to residential patients with visible confirmation of ingestion if oral. Females were divided into two groups based on their contraceptive method. The estrogen-suppressing contraceptive group included the contraceptives that suppress ovulation, which are oral contraceptives, depot medroxyprogesterone acetate (DMPA) injections, contraceptive patches, contraceptive implants, and vaginal rings. The naturally cycling group included females who were not using any contraceptives. For the current study, 65 females using IUDs were excluded, as IUDs may suppress ovulation in some women but not others (18). The final sample includes 348 females with 131(37.6%) having a BPD diagnosis and 145 (41.7%) using estrogen-suppressing contraceptives.

### Statistical Analysis

We performed multiple linear regressions to test the association of behavioral and functional difficulties at admission and discharge with co-morbid BPD diagnosis and estrogen-suppressing contraceptive use. In addition, we conducted linear mixed models with a random intercept for participants to evaluate the association of behavioral and functional difficulties in the overall sample that combines admission and discharge measures. To explore these associations longitudinally, we used linear mixed models with behavioral and functional difficulties at admission and discharge as the dependent variable, co-morbid BPD diagnosis or estrogen-suppressing contraceptives as the main effect, (co-morbid BPD diagnosis x time point) or (estrogen-suppressing contraceptives x time point) as the interaction term, and a random intercept for subjects.

Finally, we examined estrogen-suppressing contraceptive use as the moderator of the associations between behavioral and functional difficulties at admission and discharge and co-morbid BPD diagnosis. We also examined the moderation effect longitudinally, by incorporating a three-way interaction term (co-morbid BPD diagnosis x estrogen-suppressing contraceptives x time point) in the models. All analyses were performed using R Statistical Software (v4.2.1; R Core Team 2022).

## Results

### Demographic and clinical characteristics

Demographic and clinical characteristics of the analytical sample are presented in Table 1 and Supplementary Table 1. Participants with a co-morbid BPD diagnosis are more likely to be younger and have a primary diagnosis of a mood disorder (e.g., bipolar disorder, major depressive disorder; Table 1). Females using estrogen-suppressing contraceptives are also more likely to be younger than naturally cycling females (Supplementary Table 1).

**Table 1:**
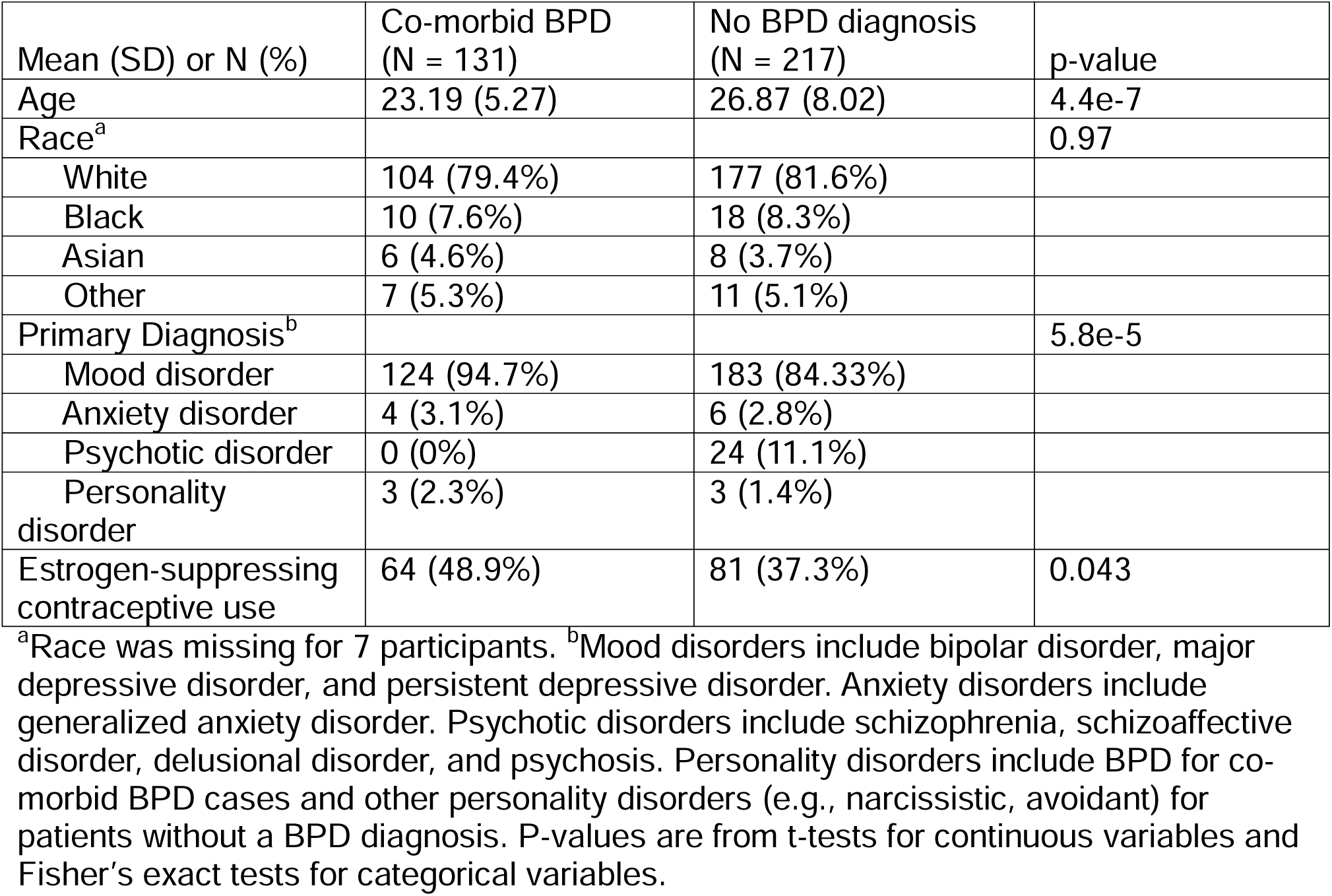
Demographic and clinical characteristics.

| Mean (SD) or N (%) | Co-morbid BPD<br>(N = 131) | No BPD diagnosis<br>(N = 217) | p-value |
| --- | --- | --- | --- |
| Age | 23.19 (5.27) | 26.87 (8.02) | 4.4e-7 |
| Race<sup>a</sup> |  |  | 0.97 |
| White | 104 (79.4%) | 177 (81.6%) |  |
| Black | 10 (7.6%) | 18 (8.3%) |  |
| Asian | 6 (4.6%) | 8 (3.7%) |  |
| Other | 7 (5.3%) | 11 (5.1%) |  |
| Primary Diagnosis<sup>b</sup> |  |  | 5.8e-5 |
| Mood disorder | 124 (94.7%) | 183 (84.33%) |  |
| Anxiety disorder | 4 (3.1%) | 6 (2.8%) |  |
| Psychotic disorder | 0 (0%) | 24 (11.1%) |  |
| Personality disorder | 3 (2.3%) | 3 (1.4%) |  |
| Estrogen-suppressing contraceptive use | 64 (48.9%) | 81 (37.3%) | 0.043 |

<sup>a</sup>Race was missing for 7 participants. <sup>b</sup>Mood disorders include bipolar disorder, major depressive disorder, and persistent depressive disorder. Anxiety disorders include generalized anxiety disorder. Psychotic disorders include schizophrenia, schizoaffective disorder, delusional disorder, and psychosis. Personality disorders include BPD for co-morbid BPD cases and other personality disorders (e.g., narcissistic, avoidant) for patients without a BPD diagnosis. P-values are from t-tests for continuous variables and Fisher's exact tests for categorical variables.

### Associations between co-morbid BPD and behavioral and functional difficulties

We first examined the difference in the severity of behavioral and functional difficulties between females with and without a co-morbid BPD diagnosis. Females with a BPD diagnosis exhibited increased difficulties in relationships and impulsive behaviors in the overall sample, as well as at admission and discharge separately (Table 2, Supplementary Figure 1A and 1D). Additionally, females with a co-morbid BPD diagnosis showed a greater improvement in impulsivity (β[SE] = -0.28[0.08], p = 6.1e-4, Supplementary Figure 1D, Supplementary Table 2). However, this greater improvement associated with BPD diagnosis might be due to patients with BPD having higher impulsive behaviors to begin with (Supplementary Figure 1D). Patients with co-morbid BPD exhibited more severe depression and anxiety symptoms only at discharge (Table 2, Supplementary Figure 1C). Finally, BPD was associated with increased difficulties in daily living in the overall sample but not when examined separately at admission and discharge (Table 2). The associations were largely consistent with the same direction of effect in the sensitivity analyses adjusting for primary diagnosis (Supplementary Table 3).

**Table 2:** The associations between co-morbid BPD and behavioral and functional difficulties.

| Admission (N = 348 samples) |  |  |  |
| --- | --- | --- | --- |
| Psychiatric Symptoms | Beta ($\beta$) | SE | p-value |
| Difficulties in relationships | 0.27 | 0.11 | **0.012** |
| Difficulties in daily living | 0.21 | 0.11 | 0.060 |
| Depression and Anxiety | 0.20 | 0.11 | 0.060 |
| Impulsivity | 0.37 | 0.08 | **9.0e-6** |
| Discharge (N = 348 samples) |  |  |  |
| Psychiatric Symptoms | Beta ($\beta$) | SE | p-value |
| Difficulties in relationships | 0.24 | 0.09 | **0.010** |
| Difficulties in daily living | 0.15 | 0.10 | 0.126 |
| Depression and Anxiety | 0.23 | 0.10 | **0.025** |
| Impulsivity | 0.13 | 0.05 | **0.017** |
| Longitudinal analysis* (N = 696 samples) |  |  |  |
| Psychiatric Symptoms | Beta ($\beta$) | SE | p-value |
| Difficulties in relationships | 0.28 | 0.10 | **0.005** |
| Difficulties in daily living | 0.21 | 0.10 | **0.038** |
| Depression and Anxiety | 0.20 | 0.10 | 0.059 |
| Impulsivity | 0.39 | 0.07 | **1.9e-8** |

\*Longitudinal analysis includes data from admission and discharge, and the associations were tested using linear mixed models with a random intercept for participants.
The models were adjusted for age. Significant associations were shown in bold. SE: Standard error.

### Estrogen-suppressing contraceptive use and behavioral and functional difficulties

We next evaluated the association of estrogen-suppressing contraceptive use with co-morbid BPD diagnosis behavioral and functional difficulties. Females with a BPD diagnosis are more likely to use estrogen-suppressing contraceptives (OR[95% CI] = 1.60[1.01, 2.55], p = 0.043), but the association did not remain significant after adjusting for age and primary diagnosis (β[SE] = 0.37[0.24], p = 0.12). Estrogen-suppressing contraceptive use was not associated with behavioral and functional difficulties at admission or discharge, and there was no change in symptom severity pre- to post-treatment (Supplementary Table 4).

### The impact of estrogen-suppressing contraceptives on behavioral and functional difficulties in BPD

Finally, we investigated the influence of estrogen-suppressing contraceptives on the association between BPD diagnosis and behavioral and functional difficulties. Estrogen-suppressing contraceptive use moderated the association between BPD and difficulties in relationships both at admission and discharge, and in the overall sample (Table 3, Figure 1A), but did not influence the amelioration of the other symptoms in those with and without a BPD diagnosis (Supplementary Tables 5 and 6). Patients with a BPD diagnosis reported more difficulties in relationships at admission and discharge, as well as in the overall sample, only if they are naturally cycling (i.e., not using estrogen-suppressing contraceptives, Table 4, Figure 1A).

**Figure 1:**
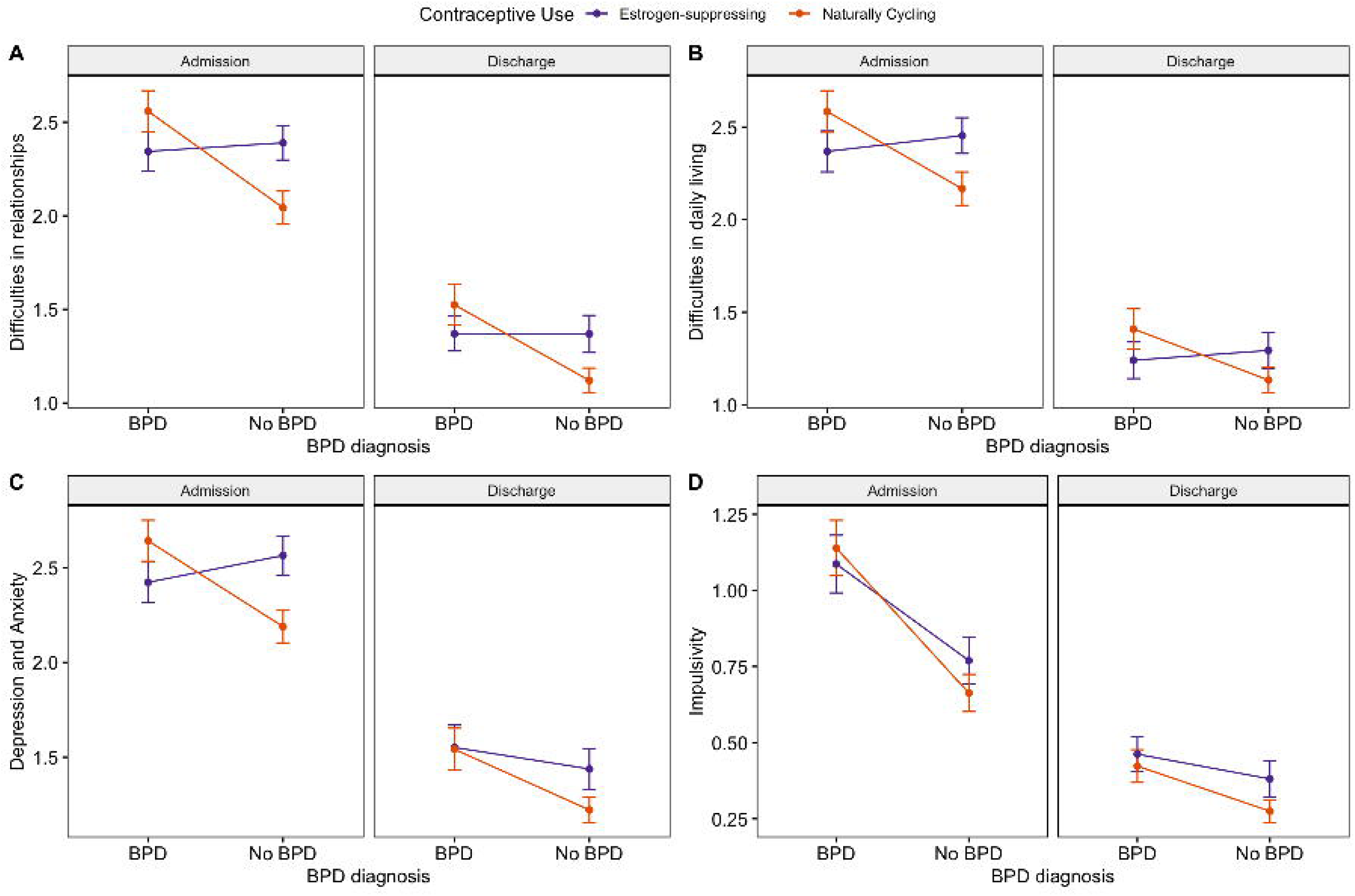
The associations between co-morbid BPD and **A)** Difficulties in relationships, **B**) Difficulties in daily living, **C)** Depression and anxiety symptoms, and **D)** Impulsivity in estrogen-suppressing contraceptive users and naturally cycling females at admission and discharge. Significant moderation effects of estrogen-suppressing contraceptives were indicated by an asterisk (*). Estrogen-suppressing contraceptives moderated the association of co-morbid BPD with difficulties in relationships at admission and discharge (A), difficulties in daily living at admission (B), and depression and anxiety symptoms at admission (C).

**Table 3:** Estrogen-suppressing contraceptives as the moderator of the associations between co-morbid BPD and behavioral and functional difficulties.

| Admission (N = 348 samples) |  |  |  |
| --- | --- | --- | --- |
| Psychiatric Symptoms | Beta ($\beta$) | SE | p-value |
| Difficulties in relationships | -0.56 | 0.21 | **0.008** |
| Difficulties in daily living | -0.50 | 0.21 | **0.020** |
| Depression and Anxiety | -0.59 | 0.21 | **0.005** |
| Impulsivity | -0.15 | 0.16 | 0.35 |
| Discharge (N = 348 samples) |  |  |  |
| Psychiatric Symptoms | Beta ($\beta$) | SE | p-value |
| Difficulties in relationships | -0.41 | 0.18 | **0.029** |
| Difficulties in daily living | -0.33 | 0.19 | 0.080 |
| Depression and Anxiety | -0.20 | 0.20 | 0.30 |
| Impulsivity | -0.07 | 0.10 | 0.52 |
| Longitudinal analysis* (N = 696 samples) |  |  |  |
| Psychiatric Symptoms | Beta ($\beta$) | SE | p-value |
| Difficulties in relationships | -0.56 | 0.20 | **0.004** |
| Difficulties in daily living | -0.50 | 0.20 | **0.013** |
| Depression and Anxiety | -0.59 | 0.20 | **0.004** |
| Impulsivity | -0.16 | 0.14 | 0.25 |

\*Longitudinal analysis includes data from admission and discharge, and the associations were tested using linear mixed models with a random intercept for participants.
The statistics were for the interaction term (Estrogen-suppressing contraceptives x BPD). The models were adjusted for age. Significant associations were shown in bold. SE: Standard error.

**Table 4:** The associations between co-morbid BPD and behavioral and functional difficulties in estrogen-suppressing contraceptive users and naturally cycling females.

|  | Estrogen-suppressing<br>contraceptive users<br>(N = 145) |  |  | Naturally Cycling<br>(N = 203) |  |  |
| --- | --- | --- | --- | --- | --- | --- |
| Admission |  |  |  |  |  |  |
| Psychiatric Symptoms | Beta (β) | SE | p-value | Beta (β) | SE | p-value |
| Difficulties in relationships | -0.05 | 0.14 | 0.75 | 0.51 | 0.15 | **0.001** |
| Difficulties in daily living | -0.06 | 0.15 | 0.69 | 0.41 | 0.16 | **0.009** |
| Depression and Anxiety | -0.15 | 0.15 | 0.32 | 0.47 | 0.15 | **0.002** |
| Impulsivity | 0.26 | 0.12 | **0.035** | 0.45 | 0.11 | **7.6e-5** |
| Discharge |  |  |  |  |  |  |
| Psychiatric Symptoms | Beta (β) | SE | p-value | Beta (β) | SE | p-value |
| Difficulties in relationships | 0.02 | 0.14 | 0.88 | 0.41 | 0.13 | **0.001** |
| Difficulties in daily living | -0.03 | 0.15 | 0.83 | 0.29 | 0.13 | **0.027** |
| Depression and Anxiety | 0.09 | 0.16 | 0.61 | 0.32 | 0.13 | **0.013** |
| Impulsivity | 0.08 | 0.09 | 0.35 | 0.15 | 0.07 | **0.024** |
| Longitudinal analysis* |  |  |  |  |  |  |
| Psychiatric Symptoms | Beta (β) | SE | p-value | Beta (β) | SE | p-value |
| Difficulties in relationships | -0.04 | 0.14 | 0.80 | 0.51 | 0.14 | **2.4e-4** |
| Difficulties in daily living | -0.06 | 0.15 | 0.67 | 0.42 | 0.14 | **0.003** |
| Depression and Anxiety | -0.16 | 0.16 | 0.31 | 0.46 | 0.14 | **8.8e-4** |
| Impulsivity | 0.29 | 0.11 | **0.007** | 0.47 | 0.09 | **5.2e-7** |

\*Longitudinal analysis includes data from admission and discharge, and the associations were tested using linear mixed models with a random intercept for participants.
The models were adjusted for age. Significant associations were shown in bold. SE: Standard error.

Estrogen-suppressing contraceptives moderated the association between BPD and the severity of depression and anxiety, and difficulties in daily living in the overall sample, as well as at admission, but not at discharge (Table 3, Figure 1B-C). Although BPD diagnosis was not associated with the severity of depression and anxiety symptoms in the overall sample, the severity of the symptoms was higher in patients with BPD who are naturally cycling (Table 4, Figure 1C). Similar to the difficulties in relationships, estrogen-suppressing contraceptives did not influence the reduction of either the severity of depression and anxiety symptoms or difficulties in daily living in patients with and without co-morbid BPD (Supplementary Tables 5 and 6). The associations remained largely consistent, showing similar effects in the sensitivity analyses that adjusted for primary diagnosis (Supplementary Tables 7 and 8).

The association between a co-morbid BPD diagnosis and higher impulsivity was consistent in patients using estrogen-suppressing contraceptives and naturally cycling patients (Table 4), and this association was not moderated by estrogen-suppressing contraceptives (Table 3, Figure 1D). When stratified by estrogen-suppressing contraceptive use, the improvement in impulsivity associated with BPD was only observed in naturally cycling females (Supplementary Table 6, Supplementary Figure 2) and not moderated by the use of estrogen-suppressing contraceptives (Supplementary Table 5). The lack of a moderating effect from estrogen-suppressing contraceptives suggests that it does not alter the fundamental association between BPD and the reduction of impulsive behaviors, but it may influence the extent of this reduction, making it more pronounced in naturally cycling females.

## Discussion

Females with BPD experience severe and rapidly shifting emotional, interpersonal, and behavioral symptoms, often co-morbid with mood and anxiety disorders (1, 3). Recent evidence highlights the role of fluctuating ovarian hormones in these rapid shifts and the severity of symptoms related to BPD (2, 8–11). Here, we investigated the impact of estrogen-suppressing contraceptives on behavioral and functional difficulties in those with and without a BPD diagnosis to explore whether contraceptives that suppress ovulation, and with it the estrogen peak, may impact the manifestation and severity of psychiatric outcomes related to BPD.

In the current study, the use of estrogen-suppressing contraceptives was not associated with the severity of behavioral and functional difficulties either at admission and discharge or change in symptom severity pre- to post-treatment. Although females with a co-morbid BPD diagnosis were more likely to use estrogen-suppressing contraceptives compared to females without, the association did not remain significant after adjusting for age and primary diagnosis. This finding is consistent with an earlier study reporting higher levels of BPD symptoms in women using oral contraceptives (11). However, we cannot assess the directionality of this association with our current sample, since the participants who were on estrogen-suppressing contraceptives had them prescribed before initiating the residential treatment and we do not have any information as to whether they were prescribed estrogen-suppressing contraceptives to manage their behavioral symptoms or for other reasons (i.e., birth control, menstrual regulation).

The reports regarding whether hormonal contraceptives increase or decrease the risk of mental health disorders are mixed (12–16). However, recent evidence suggests that the association between hormonal contraceptives and mental health outcomes may depend on individuals’ psychiatric history (14). While some studies reported mood-related disturbances as a side effect of contraceptives in the general population (12, 13), oral contraceptive pills were shown to be effective in the treatment of premenstrual dysphoric disorder (PMDD) and premenstrual worsening of depressive symptoms in females with MDD (19–22). Moreover, a recent meta-analysis reported that the use of hormonal contraceptives was associated with a decrease in depressive symptoms in women with pre-existing psychiatric disorders but an increased risk of depression in women without any history of psychiatric disorders (14). Therefore, in addition to examining the associations between BPD, estrogen-suppressing contraceptives, and psychiatric outcomes, we sought to explore the interaction between estrogen-suppressing contraceptives and BPD. As expected, a co-morbid BPD diagnosis was associated with more severe behavioral and functional difficulties, specifically increased difficulties in relationships, impulsivity, depression and anxiety symptoms, and difficulties in daily living. Importantly, we showed that estrogen-suppressing contraceptives moderated the association of BPD with difficulties in relationships, depression and anxiety symptoms, and difficulties in daily living, such that the associations between BPD and the severity of psychiatric outcomes are more pronounced in naturally cycling females, compared to females using estrogen-suppressing contraceptives. However, the use of estrogen-suppressing contraceptives did not influence the associations between BPD and change in behavioral and functional difficulties during treatment. The mechanisms behind this differential impact of estrogen-suppressing contraceptives on behavioral and functional difficulties between those with and without BPD are yet to be elucidated. One potential mechanism may be through stabilizing estrogen levels, estrogen-suppressing contraceptives may reduce emotional reactivity (23). Alternatively, estrogen-suppressing contraceptives reduce testosterone levels which were shown to be elevated in those with BPD (24–26).

Although the study has numerous strengths, including a comprehensive investigation of behavioral and functional difficulties longitudinally, it is not without limitations. First, the duration of contraceptive use varied among participants, which could confound our analyses. Second, we did not perform subgroup analyses on different types of estrogen-suppressing contraceptives (i.e., combined oral contraceptives, progestin-only methods, etc.) due to power considerations. Finally, we did not account for the menstrual cycle phase and its interaction with contraceptives. We acknowledge that the influence of contraceptives on psychiatric outcomes may depend on the menstrual cycle phase (15). However, we do not have ovarian hormone measures and menstrual cycle lengths on participants to help us accurately phase their menstrual cycle. Yet, our longitudinal analyses included measures at two time points, which to some degree account for within-subject variability due to the menstrual cycle phase.

Overall, our findings suggest that estrogen-suppressing contraceptives may help to regulate the rapidly shifting emotional, interpersonal, and behavioral symptoms in those with BPD by stabilizing estrogen levels. This opens up new avenues for treatment and highlights the need for further research to confirm these results and explore additional therapeutic options.

## Data Availability

All data produced in the present study are available in a de-identified format upon reasonable request to the authors.

