## Supplemental Files for "The Impact of Estrogen-Suppressing Contraceptives on Behavioral and Functional Difficulties in Borderline Personality Disorder"

**Supplementary Table 1:** Demographic and clinical characteristics described by estrogen-suppressing birth control use

| Mean (SD) or N (%) | Estrogen-suppressing contraceptive users  (N = 145) | Naturally Cycling  (N = 203) | p-value |
| --- | --- | --- | --- |
| Age | 24.32 (6.63) | 26.31 (7.69) | 0.01 |
| Race^a^ |  |  | 0.45 |
| White | 119 (82.1%) | 162 (79.8%) |  |
| Black | 8 (5.5%) | 20 (9.9%) |  |
| Asian | 5 (3.4%) | 9 (4.4%) |  |
| Other | 9 (6.2%) | 9 (4.4%) |  |
| Primary Diagnosis^b^ |  |  | 0.30 |
| Mood disorder | 133 (91.7%) | 174 (85.7%) |  |
| Anxiety disorder | 2 (1.4%) | 8 (3.9%) |  |
| Psychotic disorder | 7 (4.8%) | 17 (8.4%) |  |
| Personality disorder | 3 (2.1%) | 3 (8.4%) |  |

^a^Race was missing for 7 participants. ^b^Mood disorders include bipolar disorder, major depressive disorder, and persistent depressive disorder. Anxiety disorders include generalized anxiety disorder. Psychotic disorders include schizophrenia, schizoaffective disorder, delusional disorder, and psychosis. Personality disorders include BPD for co-morbid BPD cases and other personality disorders (e.g., narcissistic, avoidant) for patients without a BPD diagnosis. P-values are from t-tests for continuous variables and Fisher’s exact tests for categorical variables.

**Supplementary Table 2:** The associations between co-morbid BPD and change in behavioral and functional difficulties

| Psychiatric Symptoms | Beta (β) | SE | p-value |
| --- | --- | --- | --- |
| Difficulties in relationships | -0.04 | 0.12 | 0.704 |
| Difficulties in daily living | -0.07 | 0.11 | 0.54 |
| Depression and Anxiety | 0.04 | 0.11 | 0.74 |
| Impulsivity | -0.28 | 0.08 | **6.1e-4** |

The statistics were for the interaction term (BPD x Time Point). Significant associations were shown in bold. SE: Standard error.

**Supplementary Table 3:** Sensitivity analyses for the associations between co-morbid BPD and behavioral and functional difficulties

| Admission (N = 348 samples) | | | |
| --- | --- | --- | --- |
| Psychiatric Symptoms | Beta (β) | SE | p-value |
| Difficulties in relationships | 0.19 | 0.11 | 0.082 |
| Difficulties in daily living | 0.13 | 0.11 | 0.24 |
| Depression and Anxiety | 0.12 | 0.11 | 0.28 |
| Impulsivity | 0.39 | 0.08 | **6.85e-6** |
| Discharge (N = 348 samples) | | | |
| Psychiatric Symptoms | Beta (β) | SE | p-value |
| Difficulties in relationships | 0.22 | 0.10 | **0.025** |
| Difficulties in daily living | 0.12 | 0.10 | 0.228 |
| Depression and Anxiety | 0.18 | 0.10 | 0.074 |
| Impulsivity | 0.14 | 0.05 | **0.012** |
| Longitudinal Analysis* (N = 696 samples) | | | |
| Psychiatric Symptoms | Beta (β) | SE | p-value |
| Difficulties in relationships | 0.23 | 0.16 | **0.025** |
| Difficulties in daily living | 0.16 | 0.10 | 0.12 |
| Depression and Anxiety | 0.13 | 0.10 | 0.21 |
| Impulsivity | 0.40 | 0.07 | **1.16e-8** |

* Longitudinal analysis includes data from admission and discharge, and the associations were tested using linear mixed models with a random intercept for participants.

The models were adjusted for age and primary diagnosis. Significant associations were shown in bold. SE: Standard error.

**Supplementary Table 4:** The associations between estrogen-suppressing contraceptive use and behavioral and functional difficulties

| Admission | | | |
| --- | --- | --- | --- |
| Psychiatric Symptoms | Beta (β) | SE | p-value |
| Difficulties in relationships | 0.14 | 0.10 | 0.17 |
| Difficulties in daily living | 0.11 | 0.11 | 0.31 |
| Depression and Anxiety | 0.16 | 0.10 | 0.13 |
| Impulsivity | 0.06 | 0.08 | 0.49 |
| Discharge | | | |
| Psychiatric Symptoms | Beta (β) | SE | p-value |
| Difficulties in relationships | 0.11 | 0.09 | 0.21 |
| Difficulties in daily living | 0.05 | 0.09 | 0.60 |
| Depression and Anxiety | 0.15 | 0.10 | 0.13 |
| Impulsivity | 0.09 | 0.05 | 0.08 |
| Longitudinal Analysis* | | | |
| Psychiatric Symptoms | Beta (β) | SE | p-value |
| Difficulties in relationships | -0.04 | 0.12 | 0.73 |
| Difficulties in daily living | -0.07 | 0.11 | 0.56 |
| Depression and Anxiety | -0.03 | 0.11 | 0.98 |
| Impulsivity | 0.004 | 0.08 | 0.97 |

* Longitudinal analysis includes data from admission and discharge, and the associations were tested using linear mixed models with a random intercept for participants and the statistics are for the interaction term (Estrogen-suppressing contraceptives x Time Point).

The models were adjusted for age. Significant associations were shown in bold. SE: Standard error.

**Supplementary Table 5:** Estrogen-suppressing contraceptives as the moderator of the associations between co-morbid BPD and change in behavioral and functional difficulties

| Psychiatric Symptoms | Beta (β) | SE | p-value |
| --- | --- | --- | --- |
| Difficulties in relationships | 0.16 | 0.24 | 0.51 |
| Difficulties in daily living | 0.17 | 0.23 | 0.46 |
| Depression and Anxiety | 0.39 | 0.23 | 0.09 |
| Impulsivity | 0.09 | 0.17 | 0.58 |

The statistics were for the 3-way interaction term (Estrogen-suppressing contraceptives x BPD x Time Point). The models were adjusted for age. SE: Standard error.

**Supplementary Table 6:** The associations between co-morbid BPD and change in behavioral and functional difficulties in estrogen-suppressing contraceptive users and naturally cycling females

|  | Estrogen-suppressing contraceptive users  (N = 145) | | | Naturally Cycling  (N = 203) | | |
| --- | --- | --- | --- | --- | --- | --- |
| Psychiatric Symptoms | Beta (β) | SE | p-value | Beta (β) | SE | p-value |
| Difficulties in relationships | 0.05 | 0.16 | 0.77 | -0.11 | 0.17 | 0.51 |
| Difficulties in daily living | 0.03 | 0.17 | 0.85 | -0.14 | 0.16 | 0.37 |
| Depression and Anxiety | 0.25 | 0.18 | 0.16 | -0.13 | 0.15 | 0.36 |
| Impulsivity | -0.24 | 0.13 | 0.07 | -0.33 | 0.11 | **0.002** |

The statistics were for the interaction term (BPD x Time Point). The models were adjusted for age. Significant associations were shown in bold. SE: Standard error.

**Supplementary Table 7:** Sensitivity analyses for interaction models testing the moderating effect of estrogen-suppressing contraceptives

| Admission (N = 348 samples) | | | |
| --- | --- | --- | --- |
| Psychiatric Symptoms | Beta (β) | SE | p-value |
| Difficulties in relationships | -0.51 | 0.21 | **0.015** |
| Difficulties in daily living | -0.47 | 0.22 | **0.031** |
| Depression and Anxiety | -0.56 | 0.21 | **0.008** |
| Impulsivity | -0.15 | 0.16 | 0.349 |
| Discharge (N = 348 samples) | | | |
| Psychiatric Symptoms | Beta (β) | SE | p-value |
| Difficulties in relationships | -0.38 | 0.19 | **0.041** |
| Difficulties in daily living | -0.32 | 0.19 | 0.092 |
| Depression and Anxiety | -0.18 | 0.20 | 0.356 |
| Impulsivity | -0.10 | 0.11 | 0.348 |
| Longitudinal Analysis* (N = 696 samples) | | | |
| Psychiatric Symptoms | Beta (β) | SE | p-value |
| Difficulties in relationships | -0.53 | 0.20 | **0.008** |
| Difficulties in daily living | -0.48 | 0.20 | **0.018** |
| Depression and Anxiety | -0.57 | 0.20 | **0.006** |
| Impulsivity | -0.17 | 0.14 | 0.209 |

*Longitudinal analysis includes data from admission and discharge, and the associations were tested using linear mixed models with a random intercept for participants.

The statistics were for the interaction term (Estrogen-suppressing contraceptives x BPD). The models were adjusted for age and primary diagnosis. Significant associations were shown in bold. SE: Standard error.

**Supplementary Table 8:** Sensitivity analysis for the associations between co-morbid BPD and behavioral and functional difficulties in estrogen-suppressing contraceptive users and naturally cycling females

|  | Estrogen-suppressing contraceptive users  (N = 145) | | | Naturally Cycling  (N = 203) | | |
| --- | --- | --- | --- | --- | --- | --- |
| Admission | | | | | | |
| Psychiatric Symptoms | Beta (β) | SE | p-value | Beta (β) | SE | p-value |
| Difficulties in relationships | -0.10 | 0.15 | 0.489 | 0.41 | 0.16 | **0.011** |
| Difficulties in daily living | -0.13 | 0.15 | 0.418 | 0.32 | 0.16 | 0.051 |
| Depression and Anxiety | -0.22 | 0.16 | 0.162 | 0.37 | 0.15 | **0.017** |
| Impulsivity | 0.25 | 0.13 | **0.047** | 0.44 | 0.12 | **2.2e-4** |
| Discharge | | | | | | |
| Psychiatric Symptoms | Beta (β) | SE | p-value | Beta (β) | SE | p-value |
| Difficulties in relationships | -0.007 | 0.15 | 0.960 | 0.35 | 0.13 | **0.007** |
| Difficulties in daily living | -0.06 | 0.15 | 0.706 | 0.25 | 0.13 | 0.066 |
| Depression and Anxiety | 0.04 | 0.17 | 0.799 | 0.26 | 0.13 | 0.052 |
| Impulsivity | 0.07 | 0.09 | 0.437 | 0.17 | 0.07 | **0.018** |
| Longitudinal Analysis* | | | | | | |
| Psychiatric Symptoms | Beta (β) | SE | p-value | Beta (β) | SE | p-value |
| Difficulties in relationships | -0.08 | 0.15 | 0.587 | 0.43 | 0.14 | **0.002** |
| Difficulties in daily living | -0.11 | 0.15 | 0.472 | 0.35 | 0.14 | **0.016** |
| Depression and Anxiety | -0.22 | 0.16 | 0.179 | 0.38 | 0.14 | **0.007** |
| Impulsivity | 0.28 | 0.11 | **0.009** | 0.47 | 0.09 | **1.1e-6** |

*Longitudinal analysis includes data from admission and discharge, and the associations were tested using linear mixed models with a random intercept for participants.

The models were adjusted for age and primary diagnosis. Significant associations were shown in bold. SE: Standard error.

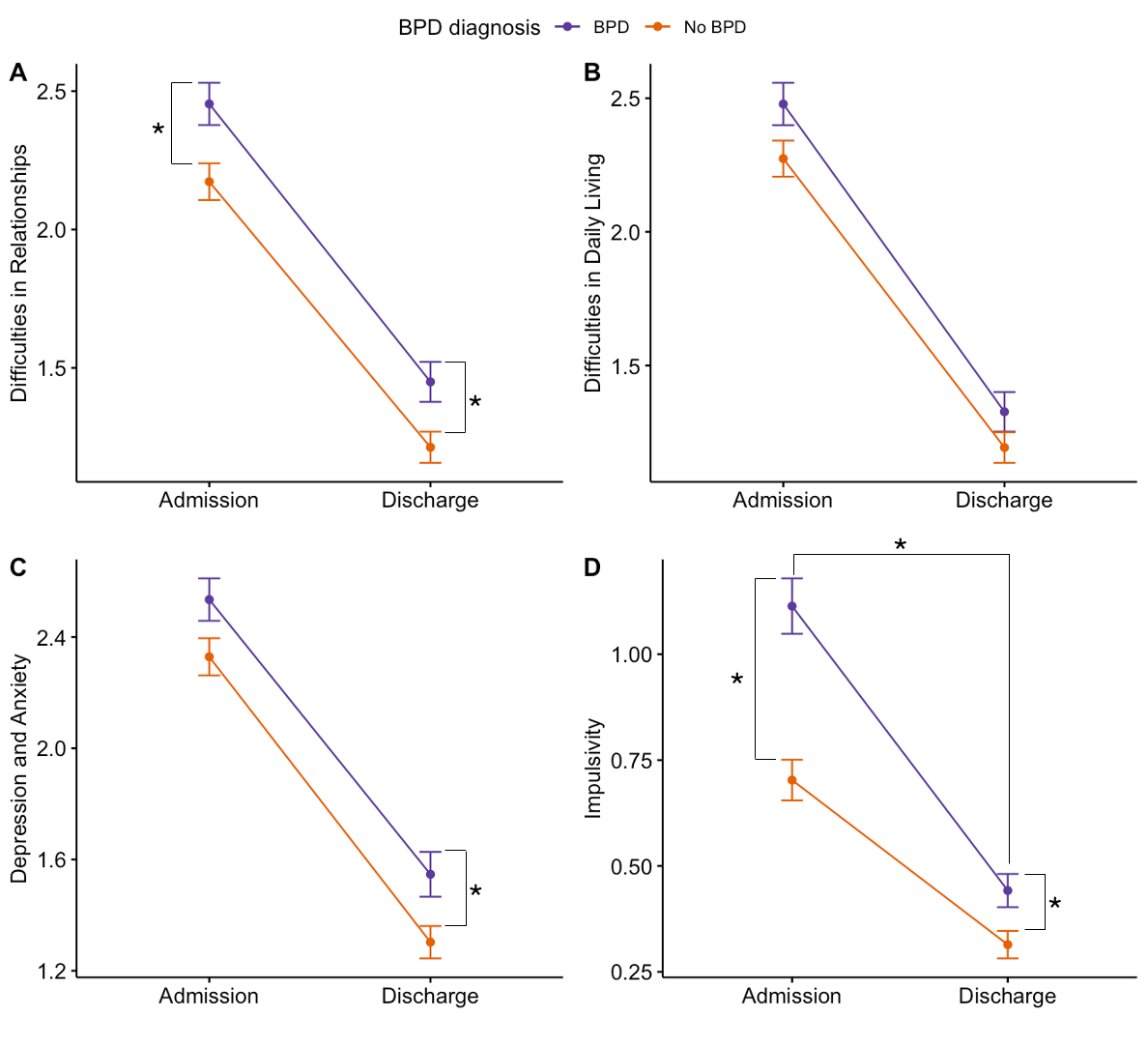

**Figure 1:** Associations between co-morbid BPD and **A)** Difficulties in relationships, **B)** Difficulties in daily living, **C)** Depression and anxiety symptoms, and **D)** Impulsivity. Significant differences between BPD and no-BPD groups in behavioral and functional difficulties at admission and discharge, and reduction of behavioral and functional difficulties were indicated by asterisk (*). BPD diagnosis was associated with increased difficulties in relationships at admission and discharge (A), increased depression symptoms at discharge (C), increased impulsivity at admission and discharge (D), and greater reduction of impulsivity pre- to post-treatment (D).

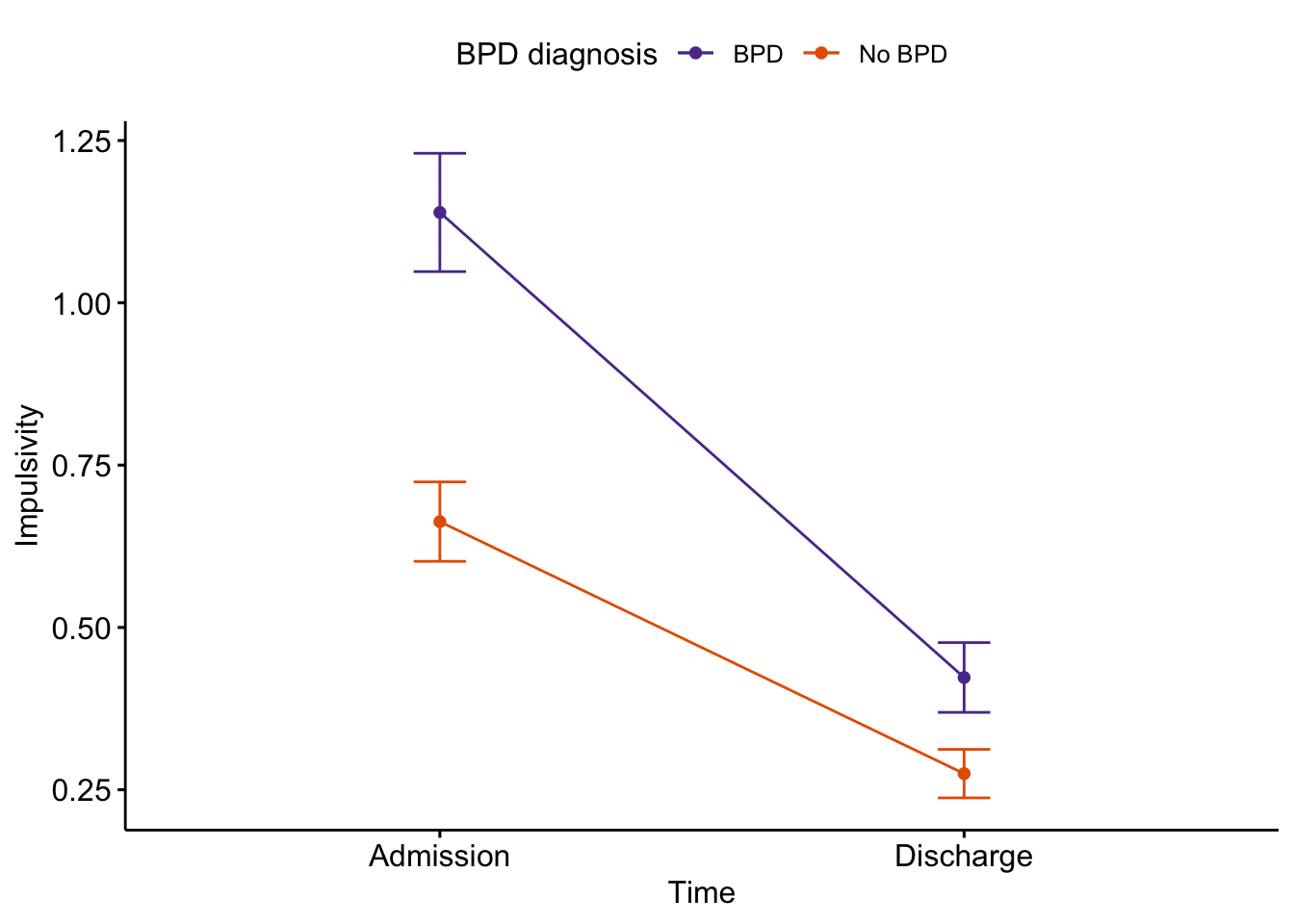

**Supplementary Figure 2:** Change in impulsive behaviors pre- to post-treatment in naturally cycling females. Naturally cycling females with co-morbid BPD showed greater improvement in impulsive behaviors compared to naturally cycling females without co-morbid BPD.
